# Unmet Needs in NSCLC Care: Insights from Temporal Routes to Treatment

**DOI:** 10.64898/2026.11.24.26368897

**Authors:** Mostafa Abbas, Neria Bitton, Rohit Sharma, Ashwini Arunachalam, Melissa L. Santorelli, Matthew A. Facktor, H. Lester Kirchner, Paul F. Simonelli, Yasser El-Manzalawy

## Abstract

**Background:** Lung cancer remains the leading cause of cancer-related deaths worldwide. Analyzing the sequence and timing of clinical events before treatment can reveal barriers to early non-small cell lung cancer (NSCLC) diagnosis and care.

**Methods:** We established a retrospective cohort of 3541 adult patients diagnosed between 2012 and 2022, and received all or part of their initial treatment within our health system. Sequences of pre-treatment clinical events—including imaging, primary care provider (PCP) and lung specialist visits, Emergency Department (ED) presentation, diagnosis, and treatment type—were encoded using 1-gram and 2-gram representations. K-means clustering, guided by the Gap Statistic method, determined the optimal pathway groups.

**Results:** Four distinct care pathways were identified: (1) Low-Dose Computed Tomography (LDCT) Screening (n=215), with high LDCT use (98.6%) and early-stage diagnoses (67.0%), yielding the highest 5-year survival rate (72.1%). (2) Moderate Early-Stage Detection (MESD) (n=1236) showed the second-highest rate of early-stage diagnoses (53.2%) with minimal LDCT uptake (0.3%) and the lowest rate of primary care engagement (15.2%). (3) High Comorbidity Burden (HCB) (n=1200), with the highest Charlson index (5), low early-stage diagnosis (36.5%), and low LDCT referral rates (3%). (4) ED Presentation (EDP) (n=890), with frequent ED visits prior to diagnosis (65.5%), low LDCT referral (1.2%), and the lowest rate of early-stage detection (19.8%).

**Conclusion:** Distinct routes revealed unmet needs, including limited LDCT screening, low primary care engagement in MESD, and barriers to early medical attention in the EDP group. Targeted interventions may enable early detection and improve survival.

## Introduction

Lung cancer is the leading cause of cancer-related deaths worldwide [1–7]. The most common type of lung cancer is (NSCLC), which is responsible for 85% of lung cancer cases [2, 8]. The late cancer stage at diagnosis limits treatment options and contributes to poor survival rates among NSCLC patients [1–4, 6, 9]. Early detection of NSCLC can significantly improve survival by identifying the disease at a localized, potentially curable stage [1, 5, 10, 11]. However, early-stage lung cancer is often asymptomatic, and when symptoms are present, they tend to be nonspecific or obscured by smoking-related conditions and other comorbid pulmonary disorders [2, 10, 12–14]. Therefore, prioritizing early detection through evidence-based screening programs and developing referral pathways is essential to improving NSCLC survival outcomes [15–17].

To improve our understanding of unmet medical needs related to cancer diagnosis, routes to diagnosis have been proposed [14, 18–23]. In their foundational work, Elliss-Brookes et al [14] defined a route to cancer diagnosis as the sequence of interactions between the patient and the health care system, which lead to a diagnosis of cancer. Briefly, they suggested seven routes, each defined using a flow diagram (i.e., pre-defined rules based on domain knowledge) and they used patients’ medical records for the six months prior to diagnosis to assign each patient to a route. Otherwise, the patient was assigned to “unknown” route. The study showed that 24% of all cancers were diagnosed through emergency presentation, which were associated with substantially lower 1-year survival rates compared to other routes. Following this work, numerous studies have adopted this framework to investigate diagnostic routes across varies international jurisdictions and patient populations [18–23].

While routes (also called pathways) to diagnosis had been studied extensively in Europe, large-scale data for US cancer patients are lacking [16]. Additionally, a key limitation of previous studies is the failure to account for frequent interactions and the temporal relationship among them. In this work we aim to establish a large cohort of NSCLC patients from a US healthcare system and address this gap using a novel approach to model the sequence of interactions between patients and the healthcare system during one year before initiation of NSCLC treatment. Our hypothesis is that a data-driven approach for identifying temporal routes to treatment in NSCLC patients will reveal novel clinically meaningful routes and enhance our understanding of the specific unmet medical needs in each route.

## Methods

### Study design

We conducted a retrospective study of electronic medical records of patients diagnosed with NSCLC at the Geisinger Health System. The Geisinger Institutional Review Board (IRB) approved the study protocol, with the approval number: 2023-1842. The need for informed consent was waived by the IRB due to the retrospective nature of the study.

### Study population

Geisinger tumor registry included 5,777 adults diagnosed with NSCLC in our health system between 2012 and 2022. **Fig. 1** shows the flow diagram of the primary study cohort selection. Patients were excluded if they had a prior history of cancer (n = 730) or lacked a documented initial treatment date (n = 854). We then excluded individuals with fewer than three clinical encounters in the year prior to initial treatment (n = 652). The final analytic cohort comprised 3,541 patients.

**Figure 1:**
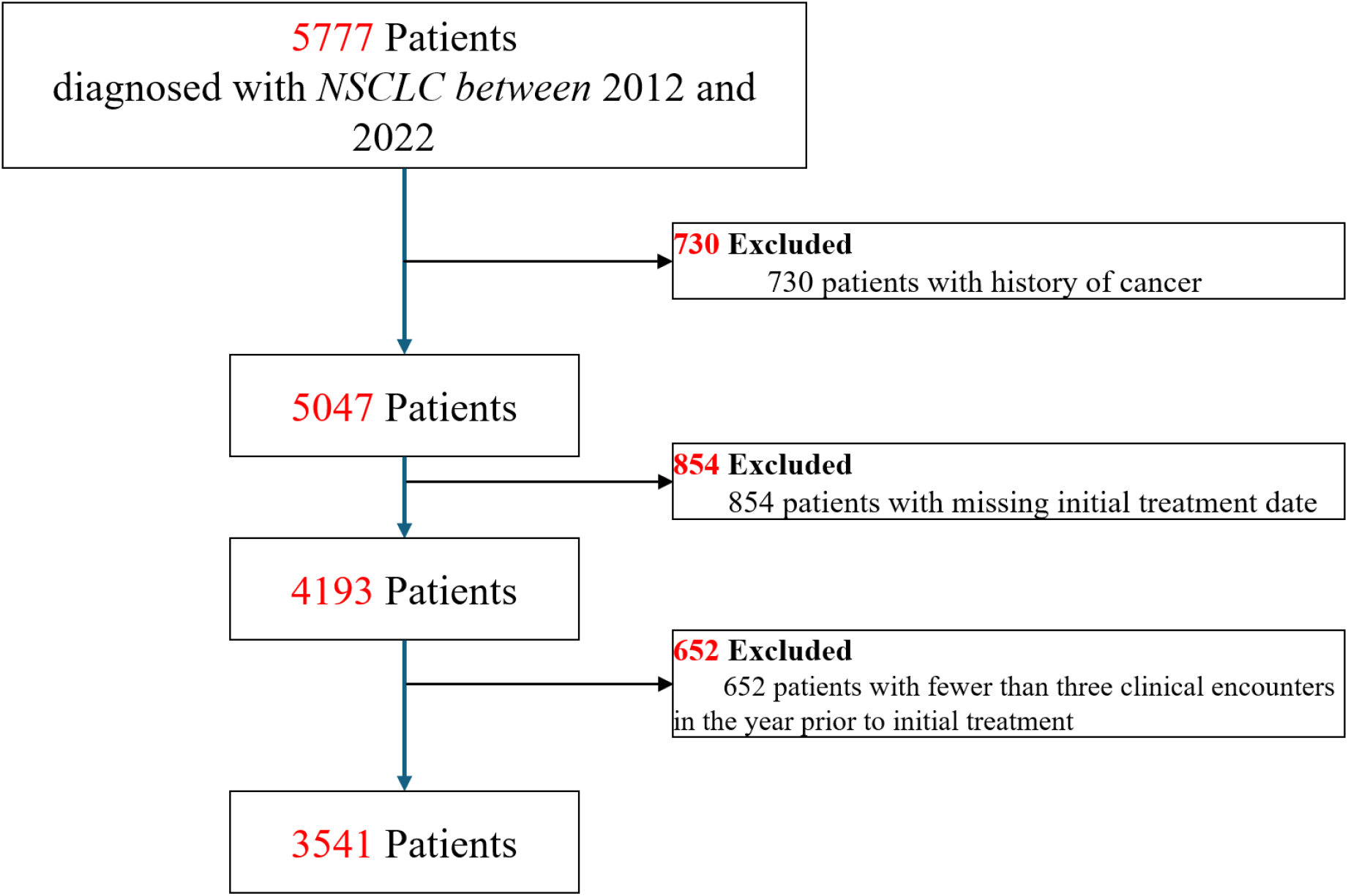
Flow diagram of the primary study cohort selection.

### Study interval and clinical events

For each patient, the study interval was defined as the 1-Year period ending on the date of initial NSCLC treatment. All clinical events occurring within this interval (including events on the treatment date) were extracted from structured EHR data including encounter records, procedure codes, radiological reports, pathology records, and tumor registry entries. **Table** 1 summarizes the clinical events used to define the temporal route to NSCLC treatment. These events included: lung specialist visit (A), primary care provider (PCP) visit (B), chest x-ray (C), chest CT (D), LDCT referral (E), LDCT examination (F), Emergency Department (ED) presentation (G), routine inpatient admission (H), biopsy procedure (I), formal NSCLC diagnosis (J), and initial treatment including the following types: chemotherapy (K), radiation therapy (L), surgery (M), and other therapies (N). All events were timestamped with their associated occurrence dates extracted from structured medical records.

**Table 1:** Clinical events occurring prior to NSCLC diagnosis and their associated codes.

| **Code** | **Event** |
|---|---|
| A | Lung specialist visit |
| B | Primary visit |
| C | Chest X-ray |
| D | Chest CT |
| E | Referral to LDCT |
| F | LDCT |
| G | ED admission |
| H | Routine admission |
| I | Biopsy |
| J | Cancer diagnosis (Dx) |
| K | Chemotherapy |
| L | Radiation Therapy |
| M | Surgery |
| N | Other initial treatment |

### Temporal routes to treatment

A patient-specific temporal route to NSCLC treatment was defined as the ordered sequence of clinical events occurring within 1-year preceding the initiation of first cancer treatment. Thus, each patient was represented as a variable-length sequence defined over a fixed alphabet of 14 encoded clinical events (**Table** 1). When multiple events occurred on the same calendar day, ties were resolved using a deterministic alphabetical ordering (A → N) based on the event codes in Table 1, ensuring reproducible sequence construction across patients. Patients were required to have a minimum of three events within the study interval, including mandatory diagnosis and treatment events, to be retained for analysis. This sequence-based representation captures both the frequency and temporal ordering of clinical events and enables identification and comparison of common routes to NSCLC treatment, as described below.

### Clustering of temporal routes to treatment

To identify common temporal routes to treatment among patients with NSCLC, we applied unsupervised machine learning-based clustering analysis [24] to the encoded clinical event sequences. Variable-length sequences were first transformed into a numeric, feature-based representation using a combination of 1-gram and 2-gram encodings [25]. The 1-gram encoding generated 14 features corresponding to the individual clinical events listed in Table 1, with each feature representing the frequency of a given event within a patient’s sequence. The 2-gram encoding captured local temporal dependencies by extracting the frequencies of all possible ordered pairs of consecutive events, yielding 14^2^ features corresponding to distinct event transitions of length two. The resulting 1-gram and 2-gram features were concatenated to form a unified feature representation for each patient. Unsupervised clustering was then performed using the *k*-means algorithm [26, 27] applied to this combined feature space. Specifically, we used the clusGap() function from the R package cluster, which applies the Gap Statistic method to determine the optimal number of clusters [28]. We used 50 Monte Carlo samples and tested 2 to 10 clusters with 25 random starts for each.

### Statistical analysis

Baseline characteristics were summarized using descriptive statistics. Categorical variables were presented as counts and percentages and compared across different groups using chi-squared test. Continuous variables were summarized as medians and interquartile ranges (IQR) and assessed for differences in distribution among groups using the Kruskal–Wallis test. Post-hoc pairwise comparisons were performed using Dunn’s test for continuous variables and chi-squared test for categorical variables.

To examine associations between clinical factors and overall survival, we constructed a multivariable Cox proportional hazards regression model using a 5-year study horizon following the initial NSCLC treatment. Survival time was measured from the date of initial treatment to death or censoring. Death events were ascertained through monthly updates to our institutional electronic health record system using linkage with the Social Security Death Index (SSDI). Patients without a documented death date were administratively censored at the earlier of 5 years (60 months) after initial treatment or the date of the final database update (November 22, 2024). Covariates included demographic factors (age and sex), smoking status, comorbidity burden (Charlson Comorbidity Index and selected individual comorbid conditions), healthcare utilization metrics (counts of primary care visits, lung specialist visits, emergency department admissions, routine inpatient admissions, and chest imaging studies), completion of LDCT screening, temporal pathway group assignment, initial treatment modality, and cancer stage at diagnosis. All numeric covariates were standardized using the z-transformation. Hazard ratios and corresponding 95% confidence intervals were estimated for all covariates.

Kaplan–Meier survival analysis was performed to characterize survival patterns across the identified temporal routes to treatment groups. All analyses were conducted in R version 4.4.2 (R Foundation for Statistical Computing, Vienna, Austria) using the survival package (version 3.7-0) for time-to-event analyses and the FSA package (version 0.10.0) for Dunn’s test, and the cluster package (version 2.1.4) for unsupervised learning.

## Results

### Baseline characteristics

**Table** 2 summarizes the baseline characteristics for the NSCLC patients in our study cohort. The study included 3541 NSCLC patients with median age 68 (interquartile range (IQR) = 61, 74) years, of whom 1663 (47%) patients were female. The median Charlson comorbidity index (CCI) was 3 (IQR = 2, 5), with chronic obstructive pulmonary disease (COPD) being the most prevalent comorbidity, affecting 38.9% of patients. A significant proportion of the patients (52.7%) were found at advanced cancer stage at the NSCLC diagnosis time, which presumably resulted in a high 5-year mortality rate of 57.8%.

**Table 2:** Patient baseline characteristics stratified by route to NSCLC treatment.

| **Variable** | **LDCT<br>(n=215)** | **MESD<br>(n=1236)** | **HCB<br>(n=1200)** | **EDP (n=890)** | **Total<br>(n=3541)** | **p-value** |
|---|---|---|---|---|---|---|
| Age | 67 (62, 72) | 68 (60, 74) | 69 (62, 76) | 67 (59, 74) | 68 (61, 74) | <0.001 |
| Female | 111 (51.6%) | 576 (46.6%) | 578 (48.2%) | 398 (44.7%) | 1663 (47%) | 0.218 |
| **Comorbidity** |  |  |  |  |  |  |
| CCI | 4 (3, 6) | 3 (2, 4) | 5 (3, 7) | 3 (2, 4) | 3 (2, 5) | <0.001 |
| Myocardial Infarction | 14 (6.5%) | 28 (2.3%) | 96 (8%) | 26 (2.9%) | 164 (4.6%) | <0.001 |
| Congestive Heart Failure | 27 (12.6%) | 62 (5%) | 188 (15.7%) | 61 (6.9%) | 338 (9.5%) | <0.001 |
| Chronic Pulmonary Disease | 128 (59.5%) | 338 (27.3%) | 720 (60%) | 190 (21.3%) | 1376 (38.9%) | <0.001 |
| Diabetes | 58 (27%) | 103 (8.3%) | 334 (27.8%) | 74 (8.3%) | 569 (16.1%) | <0.001 |
| Renal Disease | 20 (9.3%) | 56 (4.5%) | 271 (22.6%) | 47 (5.3%) | 394 (11.1%) | <0.001 |
| Smoking Status |  |  |  |  |  |  |
| CURRENT | 119 (55.3%) | 462 (37.4%) | 448 (37.3%) | 350 (39.5%) | 1379 (39%) | <0.001 |
| EVER | 95 (44.2%) | 646 (52.3%) | 668 (55.7%) | 463 (52.2%) | 1872 (52.9%) | <0.001 |
| NEVER | 1 (0.5%) | 127 (10.3%) | 84 (7%) | 74 (8.3%) | 286 (8.1%) | <0.001 |
| **Cancer Stage at diagnosis** |  |  |  |  |  |  |
| Stage I | 126 (58.6%) | 518 (41.9%) | 333 (27.8%) | 134 (15.1%) | 1111 (31.4%) | <0.001 |
| Stage II | 18 (8.4%) | 139 (11.2%) | 105 (8.8%) | 42 (4.7%) | 304 (8.6%) | <0.001 |
| Stage III | 32 (14.9%) | 238 (19.3%) | 224 (18.7%) | 157 (17.6%) | 651 (18.4%) | <0.001 |
| Stage IV | 25 (11.6%) | 256 (20.7%) | 449 (37.4%) | 484 (54.4%) | 1214 (34.3%) | <0.001 |
| Unknown stage | 14 (6.5%) | 85 (6.9%) | 89 (7.4%) | 73 (8.2%) | 261 (7.4%) | <0.001 |
| Advanced cancer stage | 57 (26.5%) | 494 (40%) | 673 (56.1%) | 641 (72%) | 1865 (52.7%) | <0.001 |
| Early stage | 144 (67%) | 657 (53.2%) | 438 (36.5%) | 176 (19.8%) | 1415 (40%) | <0.001 |
| **Initial treatment** |  |  |  |  |  |  |
| CHEMOTHERAPY | 24 (11.2%) | 248 (20.1%) | 258 (21.5%) | 212 (23.8%) | 742 (21%) | <0.001 |
| OTHER | 7 (3.3%) | 47 (3.8%) | 71 (5.9%) | 63 (7.1%) | 188 (5.3%) | <0.001 |
| RADIATION THERAPY | 97 (45.1%) | 390 (31.6%) | 571 (47.6%) | 520 (58.4%) | 1578 (44.6%) | <0.001 |
| SURGERY | 87 (40.5%) | 551 (44.6%) | 300 (25%) | 95 (10.7%) | 1033 (29.2%) | <0.001 |
| **Outcomes** |  |  |  |  |  |  |
| 5-year mortality | 60 (27.9%) | 572 (46.3%) | 778 (64.8%) | 636 (71.5%) | 2046 (57.8%) | <0.001 |

### Temporal routes to NSCLC treatment

Our clustering analysis NSCLC patients’ routes to treatment trajectories suggested that the optimal number of clusters, determined using the Gap Statistic method, is four (See Supplementary **Fig.** S1). Table 2 summarizes the baseline characteristics of the patients in each identified cluster (i.e., common temporal route to treatment). Additional characteristics of these routes are provided in **Tables** 3 - 5. In what follows, we describe the main characteristics of each identified route.

**Table 3:** Timing variables for the four routes to NSCLC treatment.

| Timing Variable (days) | LDCT (n=215) | MESD (n=1236) | HCB (n=1200) | EDP (n=890) | Total (n=3541) | p-value |
|---|---|---|---|---|---|---|
| Cancer Dx to initial treatment | 50 (29.5, 74.5) | 50 (32, 77) | 42 (27, 64.2) | 38 (23, 60) | 44 (27, 69) | <0.001 |
| Cancer Dx to follow-up | 1232 (816.5, 1909.5) | 1204 (488.8, 2333.2) | 700.5 (200, 1680.2) | 443 (125, 1296.5) | 868 (248, 1869) | <0.001 |
| Initial treatment to follow-up | 1192 (749, 1822.5) | 1156.5 (433.5, 2257.8) | 628.5 (161, 1612.2) | 382.5 (86.2, 1234.8) | 794 (195, 1800) | <0.001 |
| Last lung specialist visit to Dx | 16 (5.5, 37) | 12 (3, 29) | 11 (2, 31.8) | 13 (3, 36) | 13 (3, 32) | 0.100 |
| Last PCP visit to Dx | 31 (14, 68) | 44.5 (16, 124.5) | 19 (7, 49) | 33 (8.5, 114) | 23 (8, 62) | <0.001 |
| Last Xray to Dx | 0 (0, 48.2) | 1 (0, 27) | 7 (0, 37) | 1 (0, 9) | 2 (0, 24) | <0.001 |
| Last CT scan to Dx | 19.5 (1.5, 46.2) | 13 (0, 32) | 5 (0, 23) | 1 (0, 9) | 4 (0, 23) | <0.001 |
| Last referral for LDCT to Dx | 38 (14, 87) | 0 (0, 65) | 52.5 (22.8, 102.2) | 26 (6, 102.5) | 40 (14, 89.5) | 0.295 |
| Last LDCT imaging to Dx | 27 (7, 76) | 0 (0, 10.2) | 23 (0, 79) | 26 (13, 138) | 26 (6, 76) | 0.269 |
| Last ED admission to Dx | 122 (52.2, 190.8) | 44 (7.8, 150) | 17.5 (1, 110) | 1 (0, 17.5) | 6 (0, 74) | <0.001 |
| Last routine admission to Dx | 0 (0, 0) | 0 (0, 0) | 0 (0, 34.8) | 0 (0, 38) | 0 (0, 8) | <0.001 |
| Last biopsy to Dx | 0 (0, 0) | 0 (0, 0) | 0 (0, 41.8) | 0 (0, 0) | 0 (0, 0) | <0.001 |

**Table 4:** Summary of recorded events across routes to NSCLC treatment.

| **Event** | **LDCT (n=215)** | **MESD (n=1236)** | **HCB (n=1200)** | **EDP (n=890)** | **Total (n=3541)** | **p-value** |
|---|---|---|---|---|---|---|
| Has lung specialist visit | 195 (90.7%) | 1011 (81.8%) | 897 (74.8%) | 374 (42%) | 2477 (70%) | <0.001 |
| Has PCP visit | 156 (72.6%) | 188 (15.2%) | 1200 (100%) | 225 (25.3%) | 1769 (50%) | <0.001 |
| Has chest Xray | 113 (52.6%) | 697 (56.4%) | 992 (82.7%) | 810 (91%) | 2612 (73.8%) | <0.001 |
| Has chest CT scan | 70 (32.6%) | 775 (62.7%) | 996 (83%) | 780 (87.6%) | 2621 (74%) | <0.001 |
| Has Ref to LDCT | 214 (99.5%) | 3 (0.2%) | 36 (3%) | 11 (1.2%) | 264 (7.5%) | <0.001 |
| Has LDCT | 212 (98.6%) | 4 (0.3%) | 27 (2.2%) | 9 (1%) | 252 (7.1%) | <0.001 |
| Has ED admission | 32 (14.9%) | 231 (18.7%) | 597 (49.8%) | 699 (78.5%) | 1559 (44%) | <0.001 |
| Has routine admission | 211 (98.1%) | 1215 (98.3%) | 1018 (84.8%) | 600 (67.4%) | 3044 (86%) | <0.001 |
| Has biopsy | 207 (96.3%) | 1189 (96.2%) | 1068 (89%) | 787 (88.4%) | 3251 (91.8%) | <0.001 |
| Has lung specialist visit (before Dx) | 115 (53.5%) | 549 (44.4%) | 506 (42.2%) | 177 (19.9%) | 1347 (38%) | <0.001 |
| Has primary visit (before Dx) | 151 (70.2%) | 172 (13.9%) | 1162 (96.8%) | 167 (18.8%) | 1652 (46.7%) | <0.001 |
| Has chest Xray (before Dx) | 72 (33.5%) | 459 (37.1%) | 856 (71.3%) | 647 (72.7%) | 2034 (57.4%) | <0.001 |
| Has chest CT (before Dx) | 44 (20.5%) | 629 (50.9%) | 851 (70.9%) | 633 (71.1%) | 2157 (60.9%) | <0.001 |
| Has Ref to LDCT (before Dx) | 213 (99.1%) | 3 (0.2%) | 36 (3%) | 11 (1.2%) | 263 (7.4%) | <0.001 |
| Has LDCT (before Dx) | 209 (97.2%) | 4 (0.3%) | 27 (2.2%) | 9 (1%) | 249 (7%) | <0.001 |
| Has ED admission (before Dx) | 22 (10.2%) | 176 (14.2%) | 496 (41.3%) | 583 (65.5%) | 1277 (36.1%) | <0.001 |
| Has routine admission (before Dx) | 144 (67%) | 616 (49.8%) | 562 (46.8%) | 235 (26.4%) | 1557 (44%) | <0.001 |
| Has biopsy (before Dx) | 122 (56.7%) | 577 (46.7%) | 546 (45.5%) | 326 (36.6%) | 1571 (44.4%) | <0.001 |

**Table 5:**
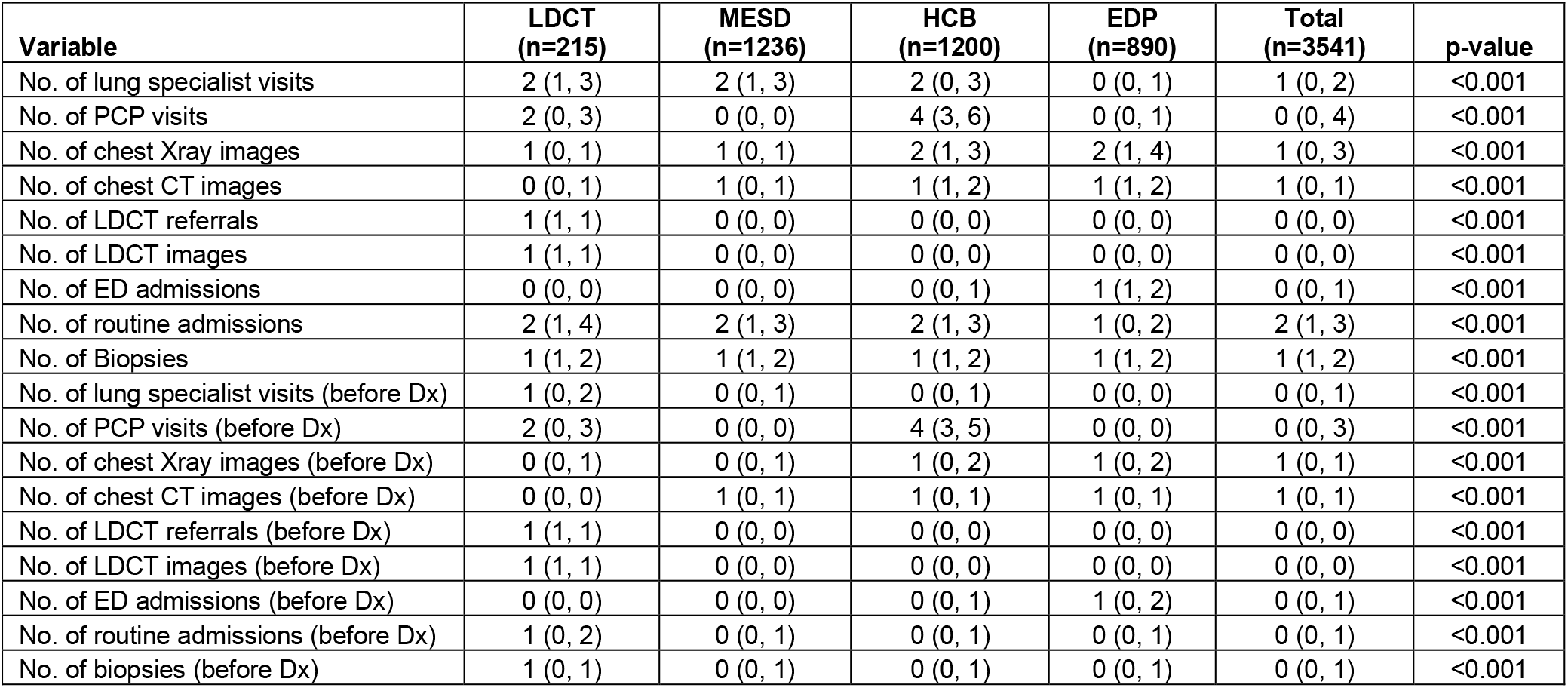
Event frequencies by route to NSCLC treatment.

| **Variable** | **LDCT<br>(n=215)** | **MESD<br>(n=1236)** | **HCB<br>(n=1200)** | **EDP<br>(n=890)** | **Total<br>(n=3541)** | **p-value** |
|---|---|---|---|---|---|---|
| No. of lung specialist visits | 2 (1, 3) | 2 (1, 3) | 2 (0, 3) | 0 (0, 1) | 1 (0, 2) | <0.001 |
| No. of PCP visits | 2 (0, 3) | 0 (0, 0) | 4 (3, 6) | 0 (0, 1) | 0 (0, 4) | <0.001 |
| No. of chest Xray images | 1 (0, 1) | 1 (0, 1) | 2 (1, 3) | 2 (1, 4) | 1 (0, 3) | <0.001 |
| No. of chest CT images | 0 (0, 1) | 1 (0, 1) | 1 (1, 2) | 1 (1, 2) | 1 (0, 1) | <0.001 |
| No. of LDCT referrals | 1 (1, 1) | 0 (0, 0) | 0 (0, 0) | 0 (0, 0) | 0 (0, 0) | <0.001 |
| No. of LDCT images | 1 (1, 1) | 0 (0, 0) | 0 (0, 0) | 0 (0, 0) | 0 (0, 0) | <0.001 |
| No. of ED admissions | 0 (0, 0) | 0 (0, 0) | 0 (0, 1) | 1 (1, 2) | 0 (0, 1) | <0.001 |
| No. of routine admissions | 2 (1, 4) | 2 (1, 3) | 2 (1, 3) | 1 (0, 2) | 2 (1, 3) | <0.001 |
| No. of Biopsies | 1 (1, 2) | 1 (1, 2) | 1 (1, 2) | 1 (1, 2) | 1 (1, 2) | <0.001 |
| No. of lung specialist visits (before Dx) | 1 (0, 2) | 0 (0, 1) | 0 (0, 1) | 0 (0, 0) | 0 (0, 1) | <0.001 |
| No. of PCP visits (before Dx) | 2 (0, 3) | 0 (0, 0) | 4 (3, 5) | 0 (0, 0) | 0 (0, 3) | <0.001 |
| No. of chest Xray images (before Dx) | 0 (0, 1) | 0 (0, 1) | 1 (0, 2) | 1 (0, 2) | 1 (0, 1) | <0.001 |
| No. of chest CT images (before Dx) | 0 (0, 0) | 1 (0, 1) | 1 (0, 1) | 1 (0, 1) | 1 (0, 1) | <0.001 |
| No. of LDCT referrals (before Dx) | 1 (1, 1) | 0 (0, 0) | 0 (0, 0) | 0 (0, 0) | 0 (0, 0) | <0.001 |
| No. of LDCT images (before Dx) | 1 (1, 1) | 0 (0, 0) | 0 (0, 0) | 0 (0, 0) | 0 (0, 0) | <0.001 |
| No. of ED admissions (before Dx) | 0 (0, 0) | 0 (0, 0) | 0 (0, 1) | 1 (0, 2) | 0 (0, 1) | <0.001 |
| No. of routine admissions (before Dx) | 1 (0, 2) | 0 (0, 1) | 0 (0, 1) | 0 (0, 1) | 0 (0, 1) | <0.001 |
| No. of biopsies (before Dx) | 1 (0, 1) | 0 (0, 1) | 0 (0, 1) | 0 (0, 1) | 0 (0, 1) | <0.001 |

#### LDCT screening route (n=215)

This was the smallest group, comprising only 6.1% of the NSCLC patients. Notably, 99.5% of the patients in this pathway had referrals for LDCT screening, and 98.6% had at least one LDCT examination (See **Table** 4). The median age of patients in this route was 67 years (IQR: 62–72), with a median CCI of 4 (IQR: 3, 6). COPD was the most prevalent comorbidity, affecting 59.5% of patients. Moreover, 55.3% of the patients were current smokers, and 67% of the patients had early cancer stage at NSCLC diagnosis date. This route was associated with the lowest 5-year mortality rate (27.9%) among the four identified common routes.

#### Moderate Early-Stage Detection (MESD) route (n=1236)

This was the largest group, encompassing 34.9% of the NSCLC patients in the study cohort. Key characteristics of this group include a median age of 68 years (IQR: 60, 74), the lowest CCI with median of 3 (IQR: 2, 4), over half of the patients (53.2%) had early NSCLC cancer stage at diagnosis. The 5-year mortality rate was 46.3%, and notably, only 15.2% of patients had documented primary care provider (PCP) visits during the study interval.

#### High Comorbidity Burden (HCB) route (n=1200)

Approximately 34% of the NSCLC patients were assigned to the HCB route. This group had the oldest median age among all groups at 69 years (IQR: 62, 76) and the highest median CCI of 5 (IQR:3, 7). This route had the highest prevalence across all five relevant comorbidities included in the baseline assessment. Notably, every patient had at least one PCP visit during the study interval, while only 74.8% had consultations with a lung specialist. The 5-year mortality rate was 64.8%, and 56.1% of the patients in this group were diagnosed with advanced cancer stage.

#### Emergency Department Presentation (EDP) route (n=890)

This group had the highest prevalence of ED presentation at 78.5%, with a median 1 day (IQR: 0, 17.5) between the last ED presentation and the cancer diagnosis date. NSCLC diagnosis following ED presentation was associated with the highest 5-year mortality rate of 71.5%. This is not surprising given that 72% of the patients were at advanced cancer stage at diagnosis time.

### Analysis of 5-year mortality after NSCLC initial treatment

Within the study cohort, the median follow-up duration was 2.2 years (IQR, 0.5–4.9 years). During the 5-year study horizon, 57.8% of patients experienced the mortality endpoint. Among patients who did not experience the event, 624 individuals (17.6%) were censored before completing 5 years of follow-up (median observed duration, 3.2 (IQR, 2.5–4.0) years, and the remaining survivors were followed for at least 5 years.

**Fig. 2** illustrates the survival curves for patients with NSCLC based on their common route to treatment. The best and poorest survivals correspond to LDCT screening and EDP routes, respectively. Notably, MESD group had better survival curve compared to the HCB group, which may be partly explained by a significantly higher rates of early stage NSCLC in the MESD group (See Table S2).

**Figure 2:**
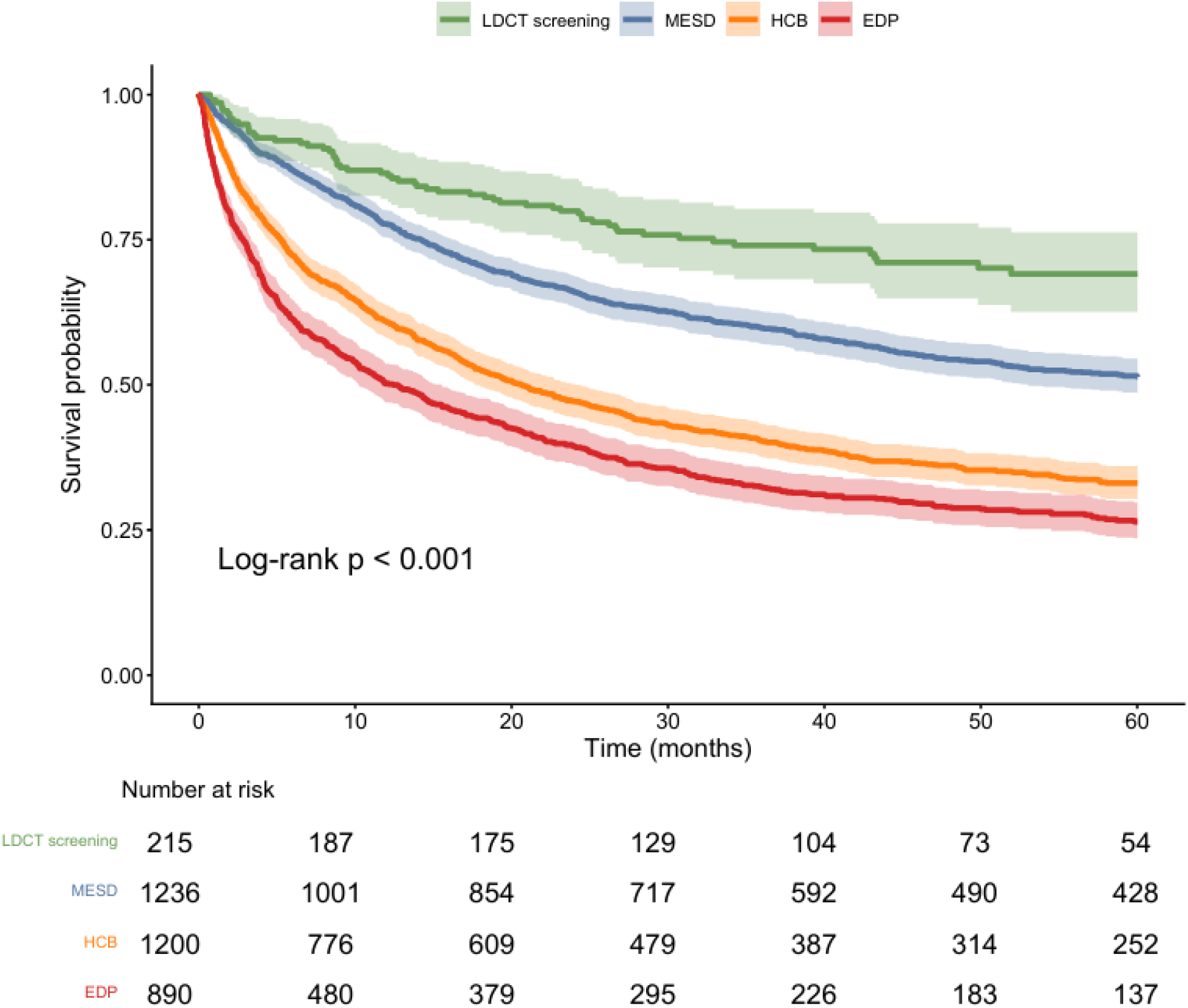
Survival curves for patients with NSCLC stratified by common routes to treatment.

**Table 6** reports the outcome of cox regression analysis of 5-year mortality indicating that significant predictors of mortality include advanced cancer stage at diagnosis (HR = 2.81, p < 0.001), advanced age (HR = 1.16, p < 0.001), assignment to EDP route (HR = 1.22, p = 0.007), and COPD (HR = 1.18, p = 0.004). Protective factors include female sex (HR = 0.82, p < 0.001), LDCT screening (HR = 0.39, p < 0.001), and surgical initial treatment (HR = 0.76, p < 0.001).

**Table 6:** Cox regression model for 5-year mortality after initial NSCLC treatment.

| **Variable** | **HR (95% CI)** | **p-value** |
|---|---|---|
| Age | 1.16 (1.09-1.24) | <0.001 |
| Female | 0.82 (0.75-0.9) | <0.001 |
| Current Smoker | 1.07 (0.97-1.17) | 0.185 |
| CCI | 1.03 (0.93-1.13) | 0.619 |
| Myocardial Infarction | 0.93 (0.75-1.14) | 0.477 |
| Congestive Heart Failure | 1.08 (0.91-1.27) | 0.391 |
| COPD | 1.18 (1.05-1.32) | 0.004 |
| Diabetes | 1.06 (0.92-1.23) | 0.42 |
| Renal Disease | 1.20 (1.01-1.42) | 0.035 |
| No. of chest X-ray | 0.99 (0.95-1.04) | 0.82 |
| No. of PCP visits | 1.09 (1.02-1.17) | 0.013 |
| No. of chest CT | 1.03 (0.97-1.08) | 0.343 |
| No. of routine admissions | 0.85 (0.8-0.9) | <0.001 |
| No. of ED admissions | 1.07 (1.03-1.11) | 0.001 |
| No. of lung specialist visits | 0.92 (0.87-0.97) | 0.004 |
| Has LDCT(Yes) | 1.40 (0.95-2.09) | 0.092 |
| **Route (reference: MESD)** |  |  |
| EDP | 1.22 (1.06-1.41) | 0.007 |
| HCB | 1.02 (0.87-1.21) | 0.778 |
| LDCT screening | 0.39 (0.24-0.63) | <0.001 |
| **Initial treatment (reference: chemotherapy)** |  |  |
| RADIATION THERAPY | 1.13 (1.01-1.26) | 0.036 |
| SURGERY | 0.76 (0.65-0.89) | <0.001 |
| Other treatment | 0.89 (0.73-1.08) | 0.24 |
| **Stage at Dx (reference: early cancer stage)** |  |  |
| Advanced cancer stage | 2.81 (2.51-3.15) | <0.001 |

### Pairwise comparisons of common temporal routes to treatment

**Tables** 2-5 demonstrate that for variables with p-value < 0.05, at least one route is significantly different from at least one of the other routes. For the completeness of our analysis, we conducted six pairwise comparisons among all possible pairs of common routes (Supplementary **Tables** S1-S6). We also conducted comparisons between each common route against other routes combined (Supplementary **Tables** S7-S10). In what follows, we highlight the most important key findings.

#### MESD vs. LDCT screening

**Supplementary Table S1.** summarizes the statistical comparisons between MESD and LDCT screening groups. Compared with the LDCT screening group, patients in the MESD group had a significantly lower CCI (3 vs. 4, p < 0.001) and lower rates of major comorbid conditions including myocardial infarction MI (2.3% vs. 6.5%, p < 0.001), congestive heart failure CHF (5.0% vs. 12.6%, p < 0.001), and chronic obstructive pulmonary disease COPD (27.3% vs. 59.5%, p < 0.001). Similarly, MESD had significantly lower rates of current smokers (37.4% vs. 55.3%, p < 0.001), lung specialist visits (81.8% vs. 90.7%, p = 0.001), and PCP visits (15.2% vs. 72.6%, p < 0.001). Although the MESD group had a significantly higher rate of chest CT examinations (62.7% vs. 32.6%, p < 0.001), these studies seem to be conducted primarily for cancer-related reasons because of the short median interval of 20 days between the first chest CT and NSCLC diagnosis.

#### HCB vs. LDCT screening

**Supplementary Table S4**. summarizes the statistical comparisons between HCB and LDCT screening groups. Several characteristics distinguished the HCB from the LDCT screening groups. For example, patients in the HCB group had significantly higher comorbidity burden in terms of higher CCI (5 vs. 4, p = 0.006) and higher rate of renal disease (22.6% vs. 9.3%, p < 0.001). Consistent with this greater comorbidity burden, the HCB group demonstrated increased healthcare utilization during the study period, including higher rates PCP visits (100% vs. 72.6%, p < 0.001), chest radiography (82.7% vs. 52.6%, p < 0.001), chest CT examinations (83.0% vs. 32.6%, p < 0.001), and ED presentations (49.8% vs. 14.9%, p < 0.001). In contrast, HCB had lower rate of current smokers (37.3% vs. 55.3%, p < 0.001) and lung specialist visits (74.8% vs. 90.7%, p < 0.001).

#### EDP vs. LDCT screening

Statistical comparisons between EDP and LDCT screening groups are presented in Supplementary Table S6. Unlike the HCB group with higher comorbidity burden compared with LDCT screening group, patients in the EDP group had significantly lower CCI (3 vs. 4, p < 0.001), rate of COPD (21.3% vs. 59.5%, p < 0.001), and prevalence of diabetes (8.3% vs. 27.0%, p < 0.001). Despite this lower comorbidity burden, the EDP group exhibited higher rates of ED presentation (78.5% vs. 14.9%, p < 0.001), chest x-ray examinations (91.0% vs. 52.6%, p < 0.001), and chest CT evaluations (87.6% vs. 32.6%, p < 0.001). Conversely, patients in the EDP group were significantly less likely to have PCP visits (25.3% vs. 72.6%, p < 0.001) or lung specialist visits (42.0% vs. 90.7%, p < 0.001).

## Discussion

Using a retrospective cohort of 3,541 patients with NSCLC, we have identified four common routes to initial NSCLC treatment. Characterizations of the cohort and the four identified routes revealed several unmet medical needs for NSCLC patients.

### Unmet needs identified in our NSCLC cohort

Our descriptive statistics (**Table** 2) highlighted three critical unmet needs within our cohort. First, there is a clear need for improved early detection. With only 40% of the NSCLC patients diagnosed at early cancer stage, it is not surprising that the five years survival rate is only 42%. Early detection of NSCLC is pivotal for improved cancer outcomes including overall survival. Second, there is a significant gap related to the underutilization of LDCT screening. Despite growing evidence linking LDCT screening to early detection and improved overall survival [1, 5, 11, 29–31], only 7.1% of the NSCLC patients in our cohort had LDCT screening. This suggests that most of the early-stage diagnoses in our cohort were made either incidentally or via symptom-driven clinical evaluation, even though early-stage NSCLC is frequently asymptomatic. Third, there is a need for reduced time from NSCLC diagnosis to initial treatment. **Table** 3 shows that the median duration from diagnosis to initial treatment is 44 days (IQR: 27, 69), indicating that more than 25% of the patients received their initial treatment more than two months after diagnosis. This exceeds the RAND-recommended benchmark of 42 days, a lung cancer-specific quality indicator to assess timeliness of care that does not depend on cancer stage at diagnosis [32].

### Unmet needs related to the LDCT screening route

Despite the advanced age, high comorbidity burden, and high prevalence of current smokers among patients in this route, it is associated with the highest rate of early-stage NSCLC diagnosis and lowest 5-year mortality rate. While adherence to LDCT screening is extremely high among patients in this route, the small proportion of patients entering it highlights an unmet need at the system level, namely improving the identification and referral of eligible populations for screening.

### Unmet needs related to the MESD route

The size of the MESD group is six times larger than that of the LDCT screening group. Compared to LDCT screening, patients in the MESD group experienced lower rates of early-stage NSCLC detection and higher five-year mortality rates. The pairwise comparison between the MESD and LDCT groups highlighted potential opportunities to improve outcomes for patients in the MESD group. First, the MESD group had a significantly lower prevalence of PCP visits (15.2% vs. 72.6%, p < 0.001), suggesting decreased engagement with routine preventative healthcare services. This limited interaction with primary care may contribute to missed opportunities for early detection and timely referral for LDCT screening. Better understanding of utilization barriers is needed as a critical step toward improving early diagnosis and survival outcomes in this group. Second, the MESD group had a significantly higher prevalence of chest CT images (62.7% vs. 32.6%, p < 0.001), suggesting potential missing opportunities for early detection of NSCLC using these opportunistic chest CT scans. Enhancing protocol and technology solutions for reviewing and following up on incidental findings in chest CTs could help bridge this gap. Third, the MESD group had significantly lower rates of current smokers (37.4% vs. 55.3%, p < 0.001) and higher rates of former smokers (52.3% vs 44.2%, p < 0.001). Former smokers with certain pack-year history are still eligible for screening, but maybe these patients are less likely to receive a referral or undergo screening. This highlights the need for improved risk-based screening strategies that go beyond smoking status to identify high-risk individuals more effectively.

### Unmet needs related to the HCB route

Patients in this group had the highest comorbidity burden among the four groups. Thus, it is not surprising that 100% had PCP visits, and approximately 75% had consultations with a lung specialist. However, only 3% of those patients were referred to LDCT screening. Despite frequent interactions with healthcare providers, these patients were rarely referred for LDCT screening. This reveals a critical gap in the implementation of lung cancer screening protocols. The low referral rate suggests that even among high-risk individuals who are regularly seen by healthcare professionals, opportunities for early detection are being overlooked (potentially due to a focus on managing existing conditions rather than prioritizing preventive care). This highlights the need for more proactive and systematic approaches to lung cancer risk assessment and early detection, particularly in high-comorbidity populations who are already embedded in the healthcare system.

### Unmet needs related to the EDP route

This pathway to NSCLC diagnosis or treatment had been extensively studied in previous research [6, 7, 16, 22, 23, 31]. Our findings showed that 25% of the NSCLC belongs to this pathway in which NSCLC diagnosis followed an ED presentation. The high proportion of diagnoses made through the ED suggests that many patients may face barriers to early medical attention, such as limited access to primary care [33], lack of awareness of symptoms [34, 35], or socioeconomic challenges [36, 37]. These findings highlight the importance of strengthening community outreach, improving access to preventive care, and implementing early risk assessment strategies to reduce reliance on emergency services for initial cancer diagnosis.

Overall, our analysis identified multiple, interconnected unmet medical needs across NSCLC routes to diagnosis and initial treatment, centered on insufficient early detection, suboptimal use of evidence-based screening, and delays in treatment initiation. Our analyses revealed that: patients diagnosed through LDCT screening route had the most favorable outcomes but remained underrepresented; those in the MESD route exhibited limited primary care engagement, underuse of opportunistic chest CT findings, and a need for more comprehensive, risk-based screening beyond age and smoking status; patients in the HCB route had frequent interaction with the healthcare system yet minimal referral to LDCT screening; and a quarter of patients were diagnosed following ED presentation, reflecting barriers to timely outpatient care and preventive services. Collectively, these findings highlight the need for systematic, risk-informed screening strategies, improved primary care engagement, better use of existing imaging (i.e., via the utilization of state-of-the-art machine learning algorithms [30, 38]), timely treatment initiation, and strengthened access to preventive and community-based care to improve NSCLC outcomes.

Routes (also called pathways) to cancer diagnosis and/or treatment have been widely studied [2, 3, 8, 10, 12–15, 17–21, 39, 40]. This work addresses two important limitations in existing literature. First, few prior studies have examined diagnostic and treatment routes (i.e., pathways) using large, real-world U.S. patient populations [6, 14, 16]. Second, most existing approaches have not explicitly accounted for the frequency and sequencing of patient interactions with the healthcare system [10, 14, 40]. By addressing these gaps, our analysis identified two novel NSCLC routes to treatment, MESD and HCB, in addition to the two widely recognized routes of screening and ED presentation. Characterization of these routes revealed distinct clinical and healthcare utilization patterns and enabled the identification of multiple, route-specific unmet medical needs across NSCLC subgroups. Despite these strengths, several limitations should be acknowledged. The use of data from a single healthcare system may limit generalizability, and route classification may be influenced by incomplete capture of healthcare encounters. Future work should prioritize external validation of these routes and assess their utility for post-diagnostic risk stratification and the design of route-specific interventions to address identified gaps in care.

## Conclusion

Using a retrospective cohort of 3541 NSCLC patients, our analysis revealed distinct temporal routes to NSCLC initial treatment and their associated unmet needs, including limited LDCT screening, low primary care engagement in the MESD route, and barriers to early medical attention in the EDP group. Targeted interventions to address these gaps could enable early NSCLC diagnosis and improve survival outcomes.

## Data Availability

The data from our study has not been deposited into a publicly available repository due to the confidential and sensitive nature of the data including protected health information.

## Ethics statement

This study was reviewed and approved by the Institutional Review Board at Geisinger Medical Center, with the approval number: 2023-1842. The need for informed consent was waived by the IRB due the retrospective nature of the study.

## Data availability statement

The data from our study has not been deposited into a publicly available repository due to the confidential and sensitive nature of the data including protected heath information.

## References

1. Horeweg N, Van Der Aalst CM, Thunnissen E, Nackaerts K, Weenink C, Groen HJ, Lammers J-WJ, Aerts JG, Scholten ET, Van Rosmalen J: Characteristics of lung cancers detected by computer tomography screening in the randomized NELSON trial. American journal of respiratory and critical care medicine 2013, 187(8):848–854.

2. Kocher F, Lunger F, Seeber A, Amann A, Pircher A, Hilbe W, Fiegl M: Incidental diagnosis of asymptomatic non–small-cell lung cancer: a registry-based analysis. Clinical lung cancer 2016, 17(1):62–67. e61.

3. Kiss Z, Bogos K, Tamasi L, Ostoros G, Mueller V, Urban L, Bittner N, Sarosi V, Vastag A, Polanyi Z: Increase in the length of lung cancer patient pathway before first-line therapy: a 6-year nationwide analysis from Hungary. Pathology and Oncology Research 2021, 27:1610041.

4. Fernández Montejo MDP, Saghir Z, Bødtger U, Jepsen R, Lynge E, Lophaven S: Identifying the population to be targeted in a lung cancer screening programme in Denmark. BMJ Open Respiratory Research 2024, 11(1):e002499.

5. Oudkerk M, Liu S, Heuvelmans MA, Walter JE, Field JK: Lung cancer LDCT screening and mortality reduction—evidence, pitfalls and future perspectives. Nature reviews Clinical oncology 2021, 18(3):135–151.

6. Pettit NR, Noriega A, Vander Missen MR: Retrospective review of patients with lung cancer presenting emergently. The American Journal of Emergency Medicine 2023, 71:129–133.

7. Venning B, Lei S, Pearce A, Lee A, Emery JD: Diagnostic testing patterns for lung cancer-related symptoms in primary care: a retrospective cohort study. BMJ Open Respiratory Research 2025, 12(1).

8. Gildea TR, DaCosta Byfield S, Hogarth DK, Wilson DS, Quinn CC: A retrospective analysis of delays in the diagnosis of lung cancer and associated costs. ClinicoEconomics and Outcomes Research 2017:261–269.

9. Jacobs C, van Ginneken B: Google’s lung cancer AI: a promising tool that needs further validation. Nature Reviews Clinical Oncology 2019, 16(9):532–533.

10. Purdie S, Creighton N, White KM, Baker D, Ewald D, Lee CK, Lyon A, Man J, Michail D, Miller AA: Pathways to diagnosis of non-small cell lung cancer: a descriptive cohort study. NPJ primary care respiratory medicine 2019, 29(1):2.

11. Force UPST, Krist AH, Davidson KW, Mangione CM, Barry MJ, Cabana M, Caughey AB, Davis EM, Donahue KE, Doubeni CA: Screening for lung cancer: US Preventive Services Task Force recommendation statement. Jama 2021, 325(10):962–970.

12. Koo MM, Rubin G, McPhail S, Lyratzopoulos G: Incidentally diagnosed cancer and commonly preceding clinical scenarios: a cross-sectional descriptive analysis of English audit data. BMJ open 2019, 9(9):e028362.

13. Borg M, Hilberg O, Andersen M, Weinreich U, Rasmussen T: Increased use of computed tomography in Denmark: stage shift toward early stage lung cancer through incidental findings. Acta oncologica 2022, 61(10):1256–1262.

14. Elliss-Brookes L, McPhail S, Ives A, Greenslade M, Shelton J, Hiom S, Richards M: Routes to diagnosis for cancer–determining the patient journey using multiple routine data sets. British journal of cancer 2012, 107(8):1220–1226.

15. Norsa L, Addo I, Shaw T, Manley S, Avery S, Delaney L, Rankin N, McGregor D, White K: Cancer care pathways mapping and dissemination toolkit: lessons learnt from cancer services in NSW, Australia. Public Health Research and Practice 2023, 33(3):e33012302.

16. Pettit N, Vander Missen M, Noriega A, Lash R: Outcomes for Emergency Presentations of Lung Cancer: A Scoping Review. The Journal of Emergency Medicine 2025, 70:50–67.

17. Zon RT, Frame JN, Neuss MN, Page RD, Wollins DS, Stranne S, Bosserman LD: American Society of Clinical Oncology policy statement on clinical pathways in oncology. Journal of Oncology Practice 2016, 12(3):261–266.

18. Weller D, Vedsted P, Anandan C, Zalounina A, Fourkala EO, Desai R, Liston W, Jensen H, Barisic A, Gavin A: An investigation of routes to cancer diagnosis in 10 international jurisdictions, as part of the International Cancer Benchmarking Partnership: survey development and implementation. BMJ open 2016, 6(7):e009641.

19. Herbert A, Abel GA, Winters S, McPhail S, Elliss-Brookes L, Lyratzopoulos G: Cancer diagnoses after emergency GP referral or A&E attendance in England: determinants and time trends in Routes to Diagnosis data, 2006–2015. British Journal of General Practice 2019.

20. Danckert B, Falborg AZ, Christensen NL, Frederiksen H, Lyratzopoulos G, McPhail S, Ryg J, Vedsted P, Thomsen LA, Jensen H: Routes to diagnosis and the association with the prognosis in patients with cancer–A nationwide register-based cohort study in Denmark. Cancer epidemiology 2021, 74:101983.

21. Martins T, Abel G, Ukoumunne OC, Mounce LT, Price S, Lyratzopoulos G, Chinegwundoh F, Hamilton W: Ethnic inequalities in routes to diagnosis of cancer: a population-based UK cohort study. British Journal of Cancer 2022, 127(5):863–871.

22. Norris RP, Fuller E, Greystoke A, Todd A, Sharp L: Routes to Diagnosis in Lung Cancer—Do Socio-Demographics Matter? An English Population-Based Study. Cancers 2025, 17(11):1874.

23. Shanmugavadivel D, Liu J-F, Ritzmann TA, Ball-Gamble A, Polanco A, Ranasinghe N, Vedhara K, Ojha S, Walker D: Quantifying diagnostic intervals and routes to diagnosis for children and young people with cancer in the UK (Childhood Cancer Diagnosis study, CCD): a population-based observational study. The Lancet Regional Health–Europe 2025, 54.

24. Jain AK, Dubes RC: Algorithms for clustering data: Prentice-Hall, Inc.; 1988.

25. EL-Manzalawy Y, Dobbs D, Honavar V: Predicting linear B-cell epitopes using string kernels. Journal of Molecular Recognition: An Interdisciplinary Journal 2008, 21(4):243–255.

26. Chong B: K-means clustering algorithm: a brief review. Academic Journal of Computing & Information Science, 4(5):37–40.

27. Hartigan JA, Wong. MA: Algorithm AS 136: A k-means clustering algorithm. Journal of the royal statistical society series c (applied statistics) 1979, 28(1):100–108.

28. Tibshirani R, Walther G, Hastie T: Estimating the number of clusters in a data set via the gap statistic. Journal of the royal statistical society: series b (statistical methodology) 2001, 63(2):411–423.

29. ten Haaf K: Evaluation of the Benefits and Harms of Lung Cancer Screening With Low-Dose Computed Tomography: A Collaborative Modeling Study for the US Preventive Services Task Force. 2021.

30. Chiu H-Y, Chao H-S, Chen Y-M: Application of artificial intelligence in lung cancer. Cancers 2022, 14(6):1370.

31. Ha NT, Kamarova S, Youens D, Ho C, Bulsara MK, Doust J, Mcrobbie D, O’Leary P, Wright C, Trevithick R: Use of CT, ED presentation and hospitalisations 12 months before and after a diagnosis of cancer in Western Australia: a population-based retrospective cohort study. BMJ open 2023, 13(10):e071052.

32. Asch SM, Kerr EA, Hamilton EG, Reifel JL, McGlynn EA: Quality of care for oncologic conditions and HIV: a review of the literature and quality indicators. In.: RAND Corporation; 2000.

33. Mitchell ED, Pickwell-Smith B, Macleod U: Risk factors for emergency presentation with lung and colorectal cancers: a systematic review. BMJ open 2015, 5(4):e006965.

34. McCutchan G, Hiscock J, Hood K, Murchie P, Neal RD, Newton G, Thomas S, Thomas AM, Brain K: Engaging high-risk groups in early lung cancer diagnosis: a qualitative study of symptom presentation and intervention preferences among the UK’s most deprived communities. BMJ open 2019, 9(5):e025902.

35. Saab MM, Noonan B, Kilty C, FitzGerald S, Collins A, Lyng Á, Kennedy U, O’Brien M, Hegarty J: Awareness and help-seeking for early signs and symptoms of lung cancer: a qualitative study with high-risk individuals. European Journal of Oncology Nursing 2021, 50:101880.

36. Redondo-Sánchez D, Petrova D, Rodríguez-Barranco M, Fernandez-Navarro P, Jimenez-Moleon JJ, Sánchez M-J: Socio-economic inequalities in lung cancer outcomes: an overview of systematic reviews. Cancers 2022, 14(2):398.

37. Exarchakou A, Rachet B, Lyratzopoulos G, Maringe C, Rubio FJ: What can hospital emergency admissions prior to cancer diagnosis tell us about socio-economic inequalities in cancer diagnosis? Evidence from population-based data in England. British Journal of Cancer 2024, 130(12):1960–1968.

38. Cellina M, Cacioppa LM, Ce M, Chiarpenello V, Costa M, Vincenzo Z, Pais D, Bausano MV, Rossini N, Bruno A: Artificial intelligence in lung cancer screening: the future is now. Cancers 2023, 15(17):4344.

39. Weller D, Menon U, Zalounina Falborg A, Jensen H, Barisic A, Knudsen AK, Bergin RJ, Brewster DH, Cairnduff V, Gavin AT: Diagnostic routes and time intervals for patients with colorectal cancer in 10 international jurisdictions; findings from a cross-sectional study from the International Cancer Benchmarking Partnership (ICBP). Bmj Open 2018, 8(11):e023870.

40. Jean C, Paillaud E, Boudou-Rouquette P, Martinez-Tapia C, Pamoukdjian F, Hagège M, Bréant S, Hassen-Khodja C, Natella P-A, Cudennec T: Hospital care trajectories of older adults with cancer and the associated clinical profiles. The Oncologist 2025, 30(3):oyae301.

