## Supplementary figures and images for "Unmet Needs in NSCLC Care: Insights from Temporal Routes to Treatment"

### Supplementary Figure S1

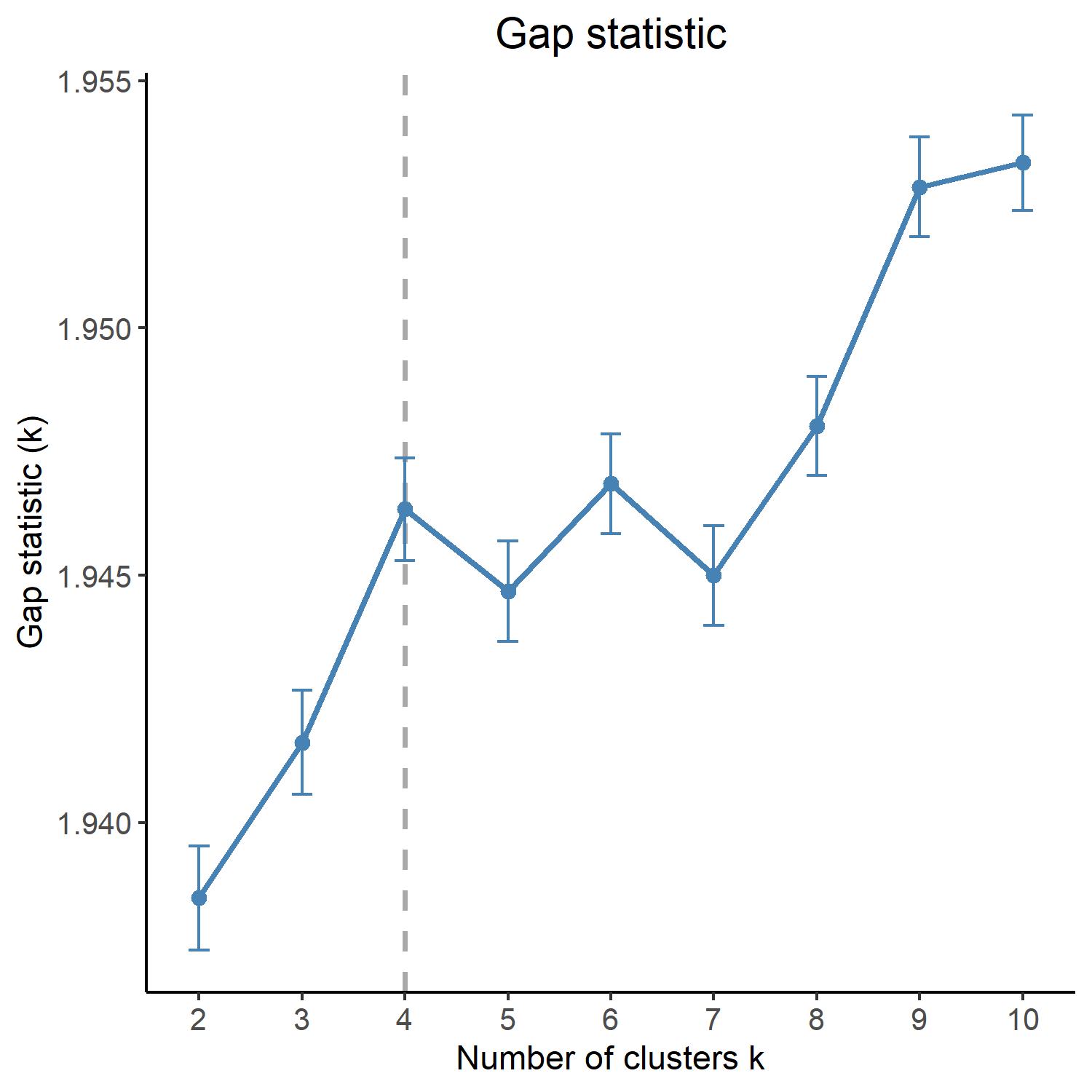
